# Association of genetic variants in enamel-formation genes with dental caries: A meta- and gene-cluster analysis

**DOI:** 10.1101/2020.09.19.20198044

**Authors:** Xueyan Li, Di Liu, Yang Sun, Jingyun Yang, Youcheng Yu

**Affiliations:** Department of Stomatology, Eye & Ent Hospital of Fudan University, Shanghai 200031, China; Beijing Key Laboratory of Clinical Epidemiology, School of Public Health, Capital Medical University, Beijing 100069, China; Department of Stomatology, Zhongshan Hospital, Fudan University, Shanghai 200032, China; Division of Statistics, School of Economics, Shanghai University, Shanghai 200444, China; Research Center of Financial Information, Shanghai University, Shanghai 200444, China; Rush Alzheimer’s Disease Center, Rush University Medical Center, Chicago, IL 60612, USA; Department of Neurological Sciences, Rush University Medical Center, Chicago, IL 60612, USA

**Keywords:** Enamel, dental caries, genetic variant, meta-analysis

## Abstract

Previous studies have reported the association between multiple genetic variants in enamel formation-related genes and the risk of dental caries with inconsistent results. We performed a systematic literature search of the PubMed, Cochrane Library, HuGE and Google Scholar databases for studies published before March 21, 2020 and conducted meta-, gene-based and gene-cluster analysis on the association between genetic variants in enamel-formation-related genes and the risk of dental caries. Our systematic literature search identified 21 relevant publications including a total of 24 studies for analysis. The genetic variant rs17878486 in *AMELX* was significantly associated with dental caries risk (OR=1.40, 95% CI: 1.02-1.93, *P*=0.037). We found no significant association between the risk of dental caries with rs12640848 in *ENAM* (OR=1.15, 95% CI: 0.88-1.52, *P*=0.310), rs1784418 in *MMP20* (OR=1.07, 95% CI: 0.76-1.49, *P*=0.702) and rs3796704 in *ENAM* (OR=1.06, 95% CI: 0.96-1.17, *P*=0.228). Gene-based analysis indicated that multiple genetic variants in *AMELX* showed joint association with the risk of dental caries (6 variants; *P*<10^−5^), so did genetic variants in *MMP13* (3 variants; *P*=0.004), *MMP2* (3 variants; *P*<10^−5^), *MMP20* (2 variants; *P*<10^−5^) and *MMP3* (2 variants; *P*<10^−5^). The gene-cluster analysis indicated a significant association between the genetic variants in this enamel-formation gene cluster and the risk of dental caries (*P*<10^−5^). The present meta-analysis revealed that genetic variant rs17878486 in *AMELX* were associated with dental caries, and multiple genetic variants in enamel-formation-related genes jointly contribute to the risk of dental caries, supporting the role of genetic variants in the enamel-formation genes in the etiology of dental caries.

## Introduction

Dental caries is one of the most common oral diseases, with an age-standardized global prevalence of untreated dentine carious lesions being around 9% in the primary dentition and around 35% in the permanent dentition during the last three decades^1^. Dental caries is a major public health concern, leading to tooth pain or loss and many other concomitants such as trouble in learning, eating or sleeping^2^. Dental caries remains to be very common despite the adoption of various preventive measures.

Dental caries results from continued localized demineralization of the dental enamel and dentine. It is a chronic disease with a multi-factorial etiology, involving the complex interactions between genetic, environmental and behavioral factors such as fluoride exposure, diet and oral hygiene^3-5^. Although previous studies have been successful in revealing the factors associated with the risk of dental caries^6-8^, more studies are needed to validate these findings, identify additional factors, and elucidate their exact roles in the etiology of dental caries.

The quality and quantity of enamel plays a direct role in the susceptibility to dental caries. Enamel formation related genes, such as *AMBN, AMELX, TUFT1, KLK4* and *ENAM*, represent a cluster of genes that are involved in the pathway of odontogenesis of dentin-containing teeth^3, 9^. Previous studies examined the association of genetic variants in this gene cluster with dental caries susceptibility, with inconsistent results^10-12^. Therefore, we performed this meta-analysis to examine the association between multiple genetic variants in this gene cluster and the risk of dental caries. Considering that the effect of individual genetic variant may be small, we also performed a gene-based and gene-cluster analysis to explore the joint association of multiple genetic variants in this gene cluster with the risk of dental caries.

## Materials and Methods

Ethical approval and informed consent statements are not required due to the systematic review and meta-analytic nature of this study.

### Eligibility Criteria

We adopted the following inclusion criteria to determine study eligibility: 1) studies on human subjects; 2) the studies has case and control group, with the case group including subjects who had caries or high caries and the control group being care-free or having low/very low caries; and 3) the studies reported data on genetic variants in enamel formation genes for subjects in both the case group and the control group. We chose the studies with a larger sample size if multiple studies used overlapping data.

Two authors (XL and JY) performed an extensive literature search of the PubMed, Cochrane Library, HuGE and Google Scholar databases for studies published before March 21, 2020. The keywords used in the literature search are provided in the online supplementary file.

We retrieved all potentially relevant publications to evaluate study eligibility. We manually searched the references in all identified studies for research that might have been missed during the literature search. We also relied on Google Scholar’s ‘cited by’ tool to search for potential publications that cited the studies identified in the literature search. The two authors performed the literature search independently. The search was limited to studies published in English. Any disagreement was resolved by group discussion (XL, SY and JY).

### Data Extraction

Two authors (DL and JY) independently extracted the following data from the eligible studies according to a pre-specified protocol for data extraction: name of the first author, year of publication, characteristics of the study participants, including sample size, mean age, distribution of gender, race/country of origin of the participants, screening method for dental caries, and genotype data for participants in the case and the control group. Any discrepancies were resolved in a group meeting. The quality of the included studies was assessed by two authors (DL and JY) independently using the Newcastle–Ottawa Scale (NOS)^13^. Extracted data were entered into a computerised spreadsheet for analysis.

### Data Analysis

We used odds ratios (ORs) as a measure of the association between genetic variants in enamel-formation genes and the risk of dental caries. In all the meta-analyses, we used random-effects models to calculate the ORs and the corresponding 95% confidence intervals (CIs). The analyses were performed using different genetic models, including additive, allelic, dominant, recessive and co-dominant genetic models. Between-study heterogeneity was assessed using I^2^, and publication bias was visually checked by a contour-enhanced funnel plot and evaluated by Egger’s test. We performed meta-analysis for a single genetic variant when there were data from multiple studies for that genetic variant. However, we only reported meta-analysis results for genetic variants that had data from at least four studies. Meta-analysis results for genetic variants that had data from less than four studies were used only in the gene-based and gene-cluster analysis as described in detail below.

### Gene-based and Gene-cluster Analysis

We conducted gene-based analyses to assess the overall association of multiple genetic variants in each enamel-formation gene with the risk of dental caries risk. We followed the methods in Dr. Li et al. 2020^14^. Specifically, we used the P-values of all genetic variants within that gene obtained from our meta-analyses or from published literature when no meta-analysis could be done. Four different P-value combination methods were utilized—the Fisher’s method^15^, the Simes method^16^, the modified inverse normal method^17^ and the modified truncated product method (TPM)^18, 19^. For the modified TPM, we calculated unweighted and weighted TPM, where the former did not consider the difference in sample sizes whereas the latter employs the sample sizes as its weight, thereby allowing studies with larger sample sizes to play a larger role in the calculation^18^. A detailed description of the four methods has been given elsewhere^19^. To estimate the P-value for the modified TPM, we ran 100,000 simulations to account for the correlation between the P-values. Gene-cluster analysis followed a similar approach as gene-based analyses.

### Sensitivity/Additional Analysis

We examined the association by including only the studies in which the genetic data in the control group satisfied Hardy–Weinberg equilibrium (HWE). We also repeated the meta-analyses by excluding studies of low quality (NOS < six stars). And finally, we examined the association through meta-analysis by including only the studies that used data from children.

All statistical analyses were performed using R (https://www.r-project.org). A P-value <0.05 was considered statistically significant. This study was reported according to the PRISMA guidelines^20^.

## Results

### Study Selection and Characteristics

**Figure 1** shows the selection of eligible studies included in our meta-analyses. We identified 132 potential publications through our initial search. After screening the abstracts, 95 publications were excluded because they were not about human subjects, were not in English, were reviews/meta-analysis or were irrelevant. This left 37 studies that were retrieved for more detailed evaluation. We excluded an additional 16 studies because they were reviews or meta-analyses, the outcomes did not include dental caries. or because there were insufficient data. This resulted in 21 publications including a total of 24 studies that met the eligibility criteria and were included in our analyses^5, 10-12, 21-38^. In summary, the meta-analyses of rs12640848 in *ENAM* included seven studies with a total of 1,256 subjects in the case group and 710 subjects in the control group^12, 25, 27, 33, 34, 37^; the meta-analyses of rs1784418 in *MMP20* included five studies with a total of 699 subjects in the case group and 817 subjects in the control group^10, 24, 26, 30, 36^; the meta-analyses of rs17878486 in *AMELX* included four studies with a total of 249 subjects in the case group and 193 subjects in the control group^10, 12, 22, 25^; and the meta-analyses of rs3796704 in *ENAM* included four studies with a total of 574 subjects in the case group and 533 subjects in the control group^12, 25, 32, 34^. Data for other genetic variants came from fewer studies.

**Figure 1.**
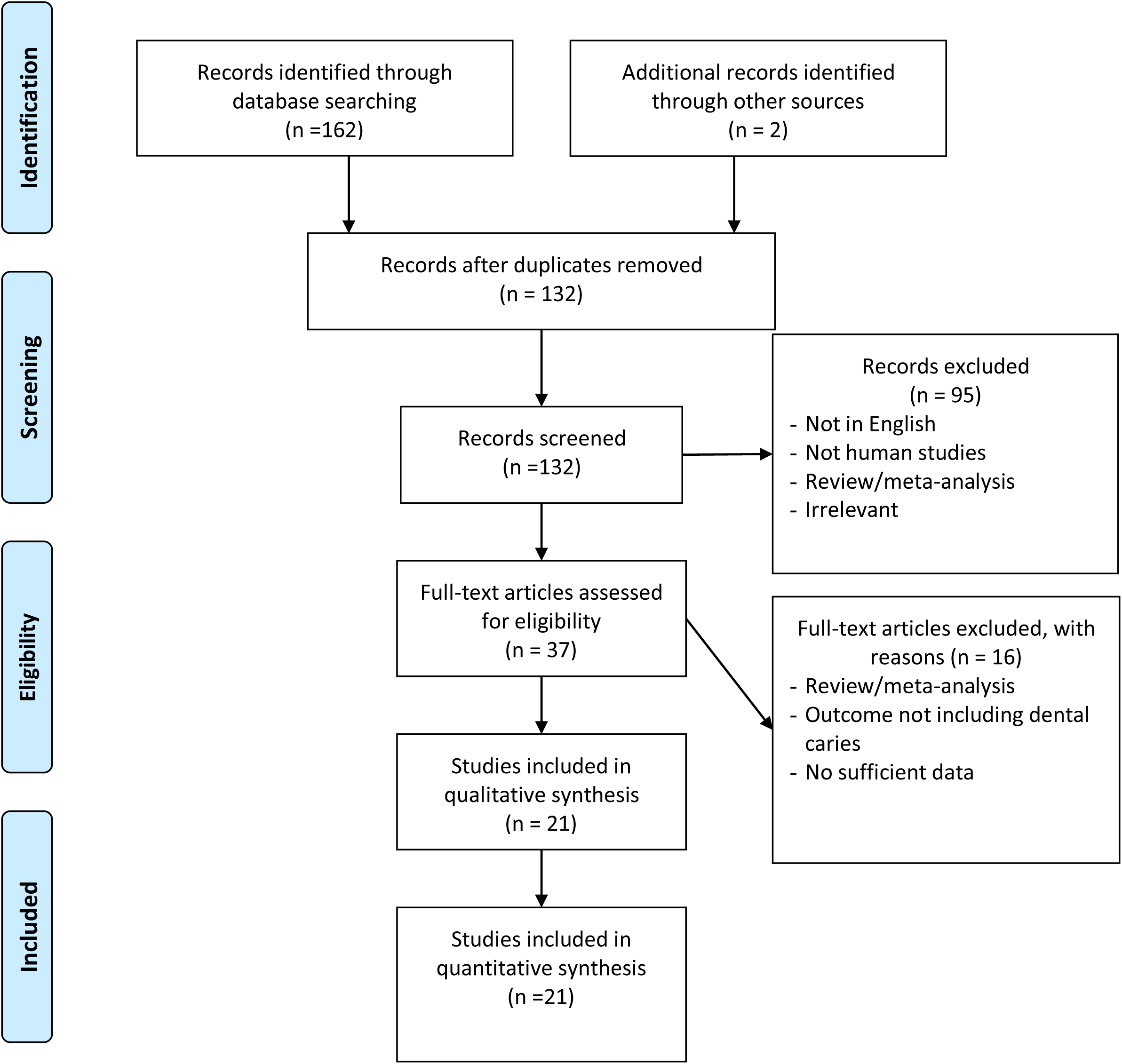
Flow diagram of the process of selecting studies included in the meta-analyses. Note: Please see the Methods section for additional details.

All included publications were published since 2008, with a sample size ranging from 71 to 1,005. The basic characteristics of the included studies are presented in **Table 1**. The majority of the included studies were of good quality, except four studies which had NOS scores of 4 or 5^5, 22, 37, 38^.

**Table 1.**
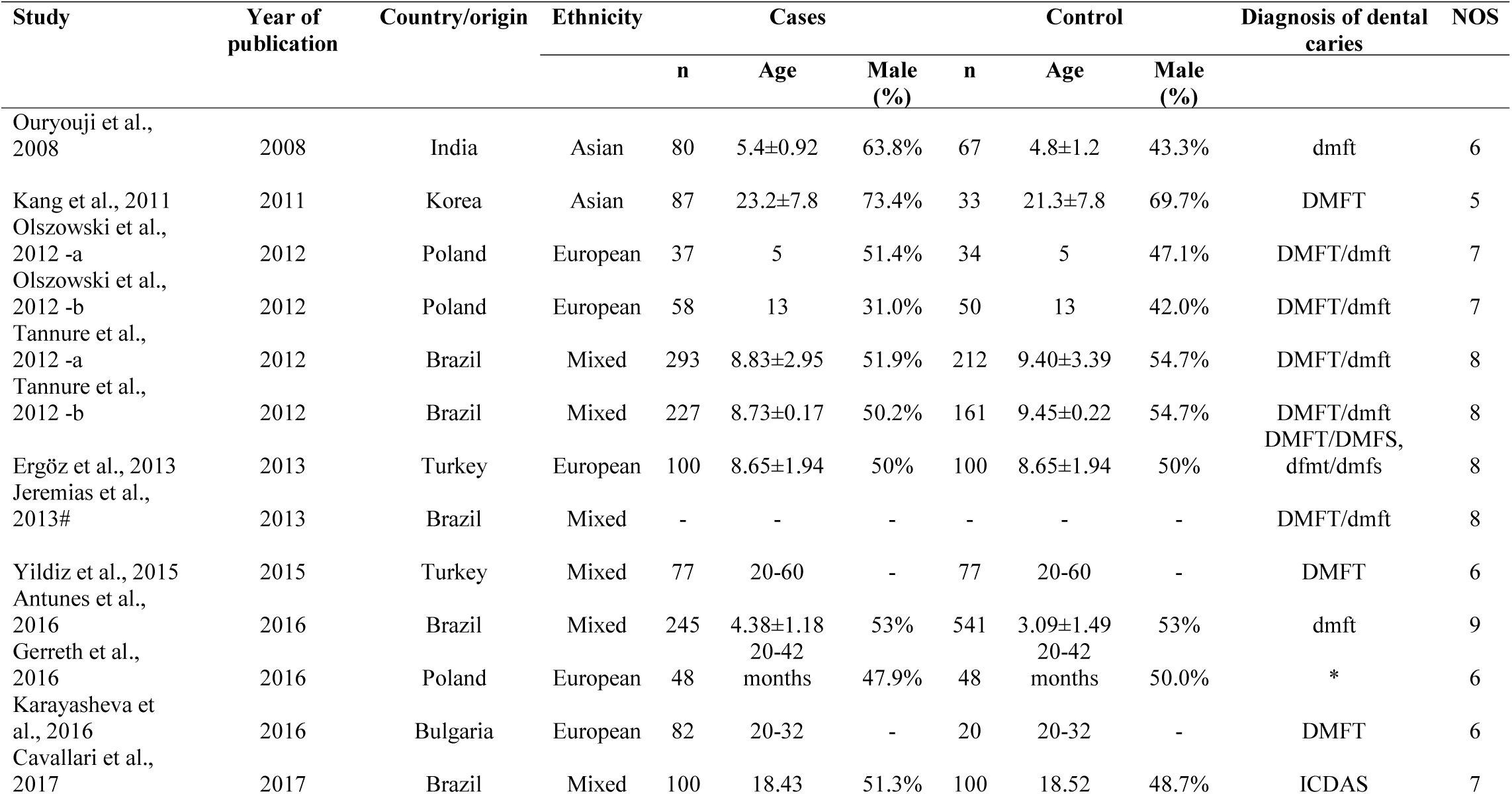
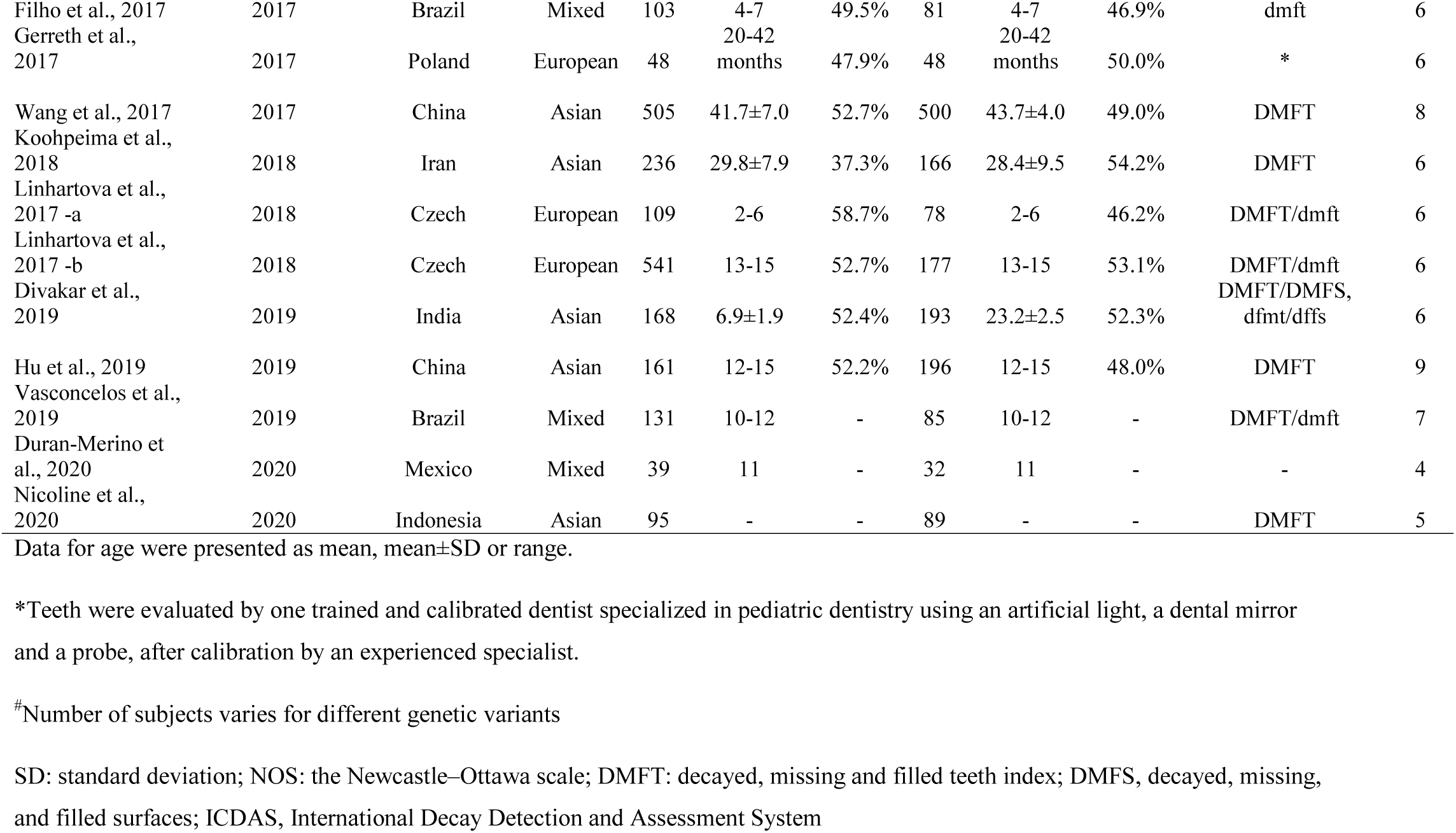
Basic characteristics of all the studies included in the analyses.

### Assessment of Publication Bias

We did not find evidence of a significant publication bias for the meta-analysis of rs12640848 in *ENAM* (*P*=0.053), rs1784418 in *MMP20* (*P*=0.238), rs17878486 in *AMELX* (P=0.521) and rs3796704 in *ENAM* (*P*=0.194; **Figure 2**). Assessment of publication bias for the meta-analysis of other genetic variants is not very meaningful due to the limited number of studies included in the corresponding meta-analysis.

**Figure 2.**
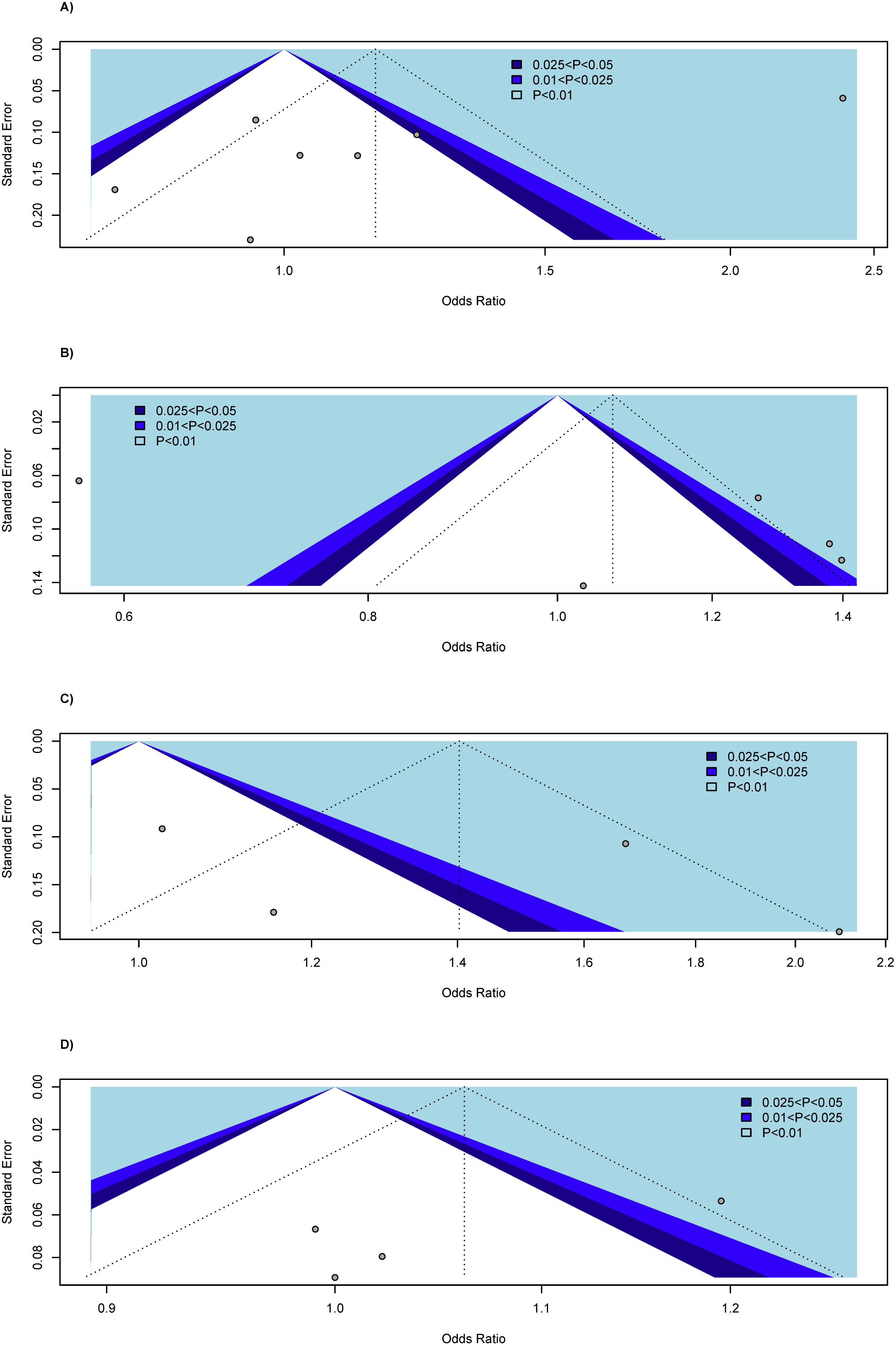
Contour-enhanced funnel plots for meta-analyses of the association with dental caries assuming an additive model.

### Association with the Risk of Dental Caries

For simplicity, we mainly reported results assuming an additive model for the meta-analyses. Complete results for meta-analyses and association analyses of individual genetic variant assuming different genetic models were presented in **Supplementary Table 1-5**. The genetic variant rs17878486 in *AMELX* was significantly associated with dental caries risk (OR=1.40, 95% CI: 1.02-1.93, *P*=0.037; **Table 2**). We found that rs12640848 in *ENAM*, the genetic variant that has largest number of studies in our meta-analysis, was not significantly associated with the risk of dental caries (OR=1.15, 95% CI: 0.88-1.52, *P*=0.310). Meta-analysis also revealed no significant association of rs1784418 (OR=1.07, 95% CI: 0.76-1.49, *P*=0.702) in *MMP20* and rs3796704 (OR=1.06, 95% CI: 0.96-1.17, *P*=0.228) in *ENAM* with the risk of dental caries. There was significant heterogeneity in the all the meta-analyses except the meta-analysis of rs3796704 (P=0.091; **Figure 3**). We did not find significant association of the four genetic variants with the risk of dental caries assuming other genetic models (**Supplementary Table 2-5)**.

**Table 2.** Meta-analysis of the association of genetic variants in enamel-formation genes with the risk of dental caries.*

| Genetic variant* | Gene | Number of studies | Number of study subjects |  | Test of heterogeneity | OR (95% CI) | P |
| --- | --- | --- | --- | --- | --- | --- | --- |
|  |  |  | Dental caries | Control |  |  |  |
| rs12640848 | *ENAM* | 7 | 1,256 | 710 | $7.69 \times 10^{-25}$ | 1.15 (0.88 - 1.52) | 0.310 |
| rs1784418 | *MMP20* | 5 | 699 | 817 | $1.38 \times 10^{-20}$ | 1.07 (0.76 - 1.49) | 0.702 |
| rs17878486 | *AMELX* | 4 | 249 | 193 | $3.58 \times 10^{-4}$ | 1.40 (1.02 - 1.93) | 0.037 |
| rs3796704 | *ENAM* | 4 | 574 | 533 | 0.091 | 1.06 (0.96 - 1.17) | 0.228 |

\*Assuming an additive genetic model

**Figure 3.**
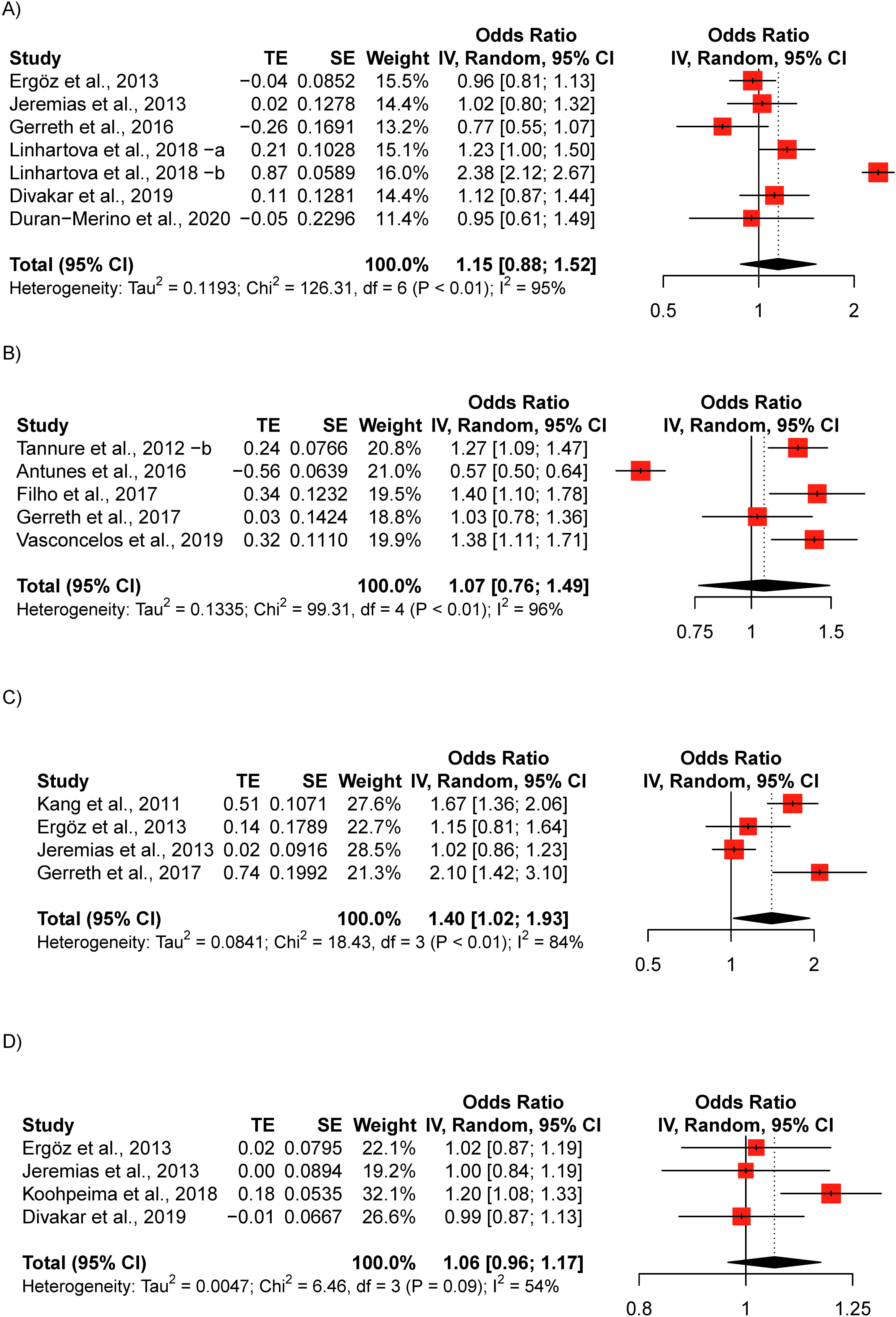
Forest plots for meta-analysis of the association with dental caries assuming an additive model. Each study is represented by a square whose area is proportional to the weight of the study. The overall effect from meta-analysis is represented by a diamond whose width represents the 95% CI for the estimated OR. A) Forest plot for meta-analysis of rs12640848; B) Forest plot for meta-analysis of rs1784418; C) Forest plot for meta-analysis of rs17878486; and D) Forest plot for meta-analysis of rs3796704 OR, odds ratio; CI, confidence interval.

Meta-analyses of other genetic variants that included fewer studies revealed no significant association with the risk of dental caries. However, multiple genetic variants in *ENAM, TUFT1, MMP2, MMP3, MMP8, MMP13, MMP20* and *AMELX* showed significant association with the risk of dental caries in individual studies (**Supplementary Table 1**).

### Gene-based and Gene-cluster Analysis

Gene-based analysis results are presented in **Table** 3. We found that multiple genetic variants in *AMELX* showed joint association with the risk of dental caries (6 variants; all Ps<2×10^−4^), so did genetic variants in *MMP13* (3 variants; all Ps≤0.01), *MMP2* (3 variants; all Ps<10^−5^), *MMP20* (2 variants; all Ps<10^−5^) and *MMP3* (2 variants; all Ps<10^− 5^). The gene-cluster analysis indicated a significant association between the genetic variants in this enamel-formation cluster and the risk of dental caries (all Ps<10^−5^; **Table 4**).

**Table 3.** Gene-based analysis of the association of genetic variants in enamel-formation genes with the risk of dental caries.

| Gene | Number of<br>variants | Fisher | Simes | Inverse | TPM<br>(unweighted) | TPM<br>(weighted) |
| --- | --- | --- | --- | --- | --- | --- |
| *ENAM* | 6 | 0.395 | 0.296 | 0.485 | 0.305 | 0.324 |
| *AMELX* | 6 | $1.26 \times 10^{-4}$ | $2.98 \times 10^{-5}$ | $1.40 \times 10^{-5}$ | $<10^{-5}$ | $<10^{-5}$ |
| *KLK4* | 5 | 0.494 | 0.276 | 0.599 | 0.275 | 0.381 |
| *TUFT1* | 4 | 0.103 | 0.077 | 0.064 | 0.090 | 0.088 |
| *MMP13* | 3 | 0.006 | 0.002 | 0.010 | 0.002 | 0.004 |
| *AMBN* | 3 | 0.452 | 0.262 | 0.998 | 0.261 | 0.199 |
| *MMP2* | 3 | $<10^{-5}$ | $4.60 \times 10^{-18}$ | $2.00 \times 10^{-51}$ | $<10^{-5}$ | $<10^{-5}$ |
| *TFIP11* | 2 | 0.514 | 0.605 | 0.459 | 0.149 | 0.150 |
| *MMP20* | 2 | $<10^{-5}$ | $3.48 \times 10^{-19}$ | $8.73 \times 10^{-18}$ | $<10^{-5}$ | $<10^{-5}$ |
| *MMP8* | 2 | $1.71 \times 10^{-4}$ | 0.005 | 0.004 | 0.004 | 0.003 |
| *MMP3* | 2 | $1.51 \times 10^{-13}$ | $1.59 \times 10^{-8}$ | $7.55 \times 10^{-9}$ | $<10^{-5}$ | $<10^{-5}$ |
| *MMP9* | 1 | 0.705 | 0.705 | 0.705 | 0.705 | 0.705 |

TPM, the modified truncated product method

**Table 4.** Gene-cluster analysis of the association of genetic variants in the enamel-formation genes with the risk of dental caries.

| Gene cluster | Fisher | Simes | Inverse | TPM |
| --- | --- | --- | --- | --- |
| Enamel-formation genes | $<10^{-5}$ | $4.17 \times 10^{-18}$ | $2.77 \times 10^{-45}$ | $<10^{-5}$ |
| TPM, the modified truncated product method |  |  |  |  |

The x-axis is the odds ratio, and the y-axis is the standard error of the estimated effect on the risk of dental caries. The vertical line in the figure represents the overall estimated odds ratio. The two diagonal lines represent the pseudo 95% confidence limits of the effect estimate.

Levels of statistical significance of the individual studies are indicated by the shaded regions: the white region corresponds to p-values greater than .05, the dark blue-shaded region corresponds to *P*-values between .025 and .05, the blue-shaded region corresponds to *P*-values between .01 and .025, and the light blue-shaded region corresponds to *P*-values below .01.

A) Funnel plot for meta-analysis of rs12640848; B) Funnel plot for meta-analysis of rs1784418;

C) Funnel plot for meta-analysis of rs17878486; and D) Funnel plot for meta-analysis of rs3796704

### Sensitivity Analysis

The association of rs17878486 in *AMELX* remained to be significant when we excluded studies in which genetic data in the control group violated HWE (OR=1.59, 95% CI: 1.15-2.20, *P*=0.006; **Supplementary Table 6**). The association disappeared after excluding studies of low quality and after excluding studies that used adult data (**Supplementary Table 7-8)**. It should be noted that the total sample sizes for these sensitivity analyses were very limited. We did not find a significant association of the other three genetic variants with the risk of dental caries in all the sensitivity analyses (**Supplementary Table 6-8)**.

## Discussions

In this manuscript, we performed a systematic literature search and conducted meta-, gene-based and gene-cluster analysis to examine the association of multiple genetic variants in the enamel formation genes with the risk of dental caries. We found that rs17878486 in *AMELX* was significantly associated with the risk of dental caries, but no significant association of other genetic variants in the meta-analyses including at least four studies. However, gene-based analysis and gene-cluster analysis indicated that genetic variants in enamel-formation-related genes were jointly associated with the risk of dental caries. To the best of our knowledge, this is the first meta-analysis on some of the genetic variants in the enamel-formation genes, and the first gene-based and gene-cluster analysis on the joint association of genetic variants in this gene cluster with the risk of dental caries.

*AMELX* is a gene located in both the X and Y chromosomes^39^ and encodes a set of isoforms of amelogenin, a major structural protein of the enamel organic matrix protein. Previous research using genetically engineered mice indicated *AMELX* was crucial for proper enamel formation^40^. Our meta-analysis of four studies including a total of 249 subjects with caries and 193 subjects without caries indicated that the genetic variant rs17878486 in *AMELX* was significantly associated with the risk of dental caries assuming an additive model; however, it showed no significant association under other genetic models (**Supplementary Table 2-5**). Moreover, no other genetic variants showed significant association except rs5933871 (**Supplementary Table 1**). Given the important role of *AMELX* in amelogenesis, future research is greatly needed to validate the relationship of the reported genetic variants and explore other genetic variants in this gene that may be associated with the risk of dental caries.

The gene *ENAM* is located on chromosome 4q 13.3 and has 10 exons. It encodes enamelin protein which is critical for the formation and elongation of enamel crystallites^41^. Meta-analysis for the genetic variant rs12640848 in *ENAM* has the largest number of included studies. Previous research suggested that this polymorphism was associated with the risk of dental caries^42, 43^. In our meta-analysis assuming an additive model, of the six publications including a total of seven studies, only one study including 541 caries-affected children and 177 caries-free children aged 13-15 years old in Czech indicated a significant positive association of the G allele with an increased risk of dental caries (OR=2.38, 95% CI: 2.12-2.67, *P*<0.0001)^33^. However, we did notice that several included studies indicated a significant association of this variant under other genetic models^27, 34^, although the overall meta-analysis results were still not significant (data not shown). Most of the other studied genetic variants in this gene were not associated with the risk of dental caries except one SNP rs3806804 which showed marginal association (OR=0.80, 95% CI: 0.65-1.00; *P*=0.049; **Supplementary Table 1**). More studies are needed to elucidate the role of rs12640848 and other genetic variants in *ENAM* in the formation of dental caries.

Matrix metalloproteinases (MMPs) refer to a large family of zinc-dependent endoproteinases which are fundamental in tooth formation and mineralization of dental tissue^44^. More than 25 vertebrate MMPs have been identified, and 24 of them are present in humans^45^. Genetic variants in many of the MMP genes, such as *MMP2, MMP3, MMP13* and *MMP20*, have been reported to be associated with the risk of dental caries^23, 24, 28^. Meta-analysis of rs1784418 in *MMP20* including a total of five studies showed no significant association of this genetic variant with the risk of dental caries (**Table 1**). However, many other genetic variants in this gene cluster were significantly associated with the risk of dental caries (**Supplementary Table 1**). These results should be interpreted with caution because they were based on data from single studies with limited sample sizes. Gene-based analysis indicated that genetic variants within *MMP2, MMP3, MMP8, MMP13* and *MMP20* jointly associated with the risk of dental caries, further supporting the involvement of genetic variants in these genes in influencing the risk of dental caries. Future studies are warranted to reveal the function of these genetic variants in the etiology of dental caries.

We observed significant heterogeneities in all of the meta-analyses except the meta-analysis for rs3796704 in *ENAM* (P=0.091; **Table 2**). The heterogeneity disappeared in some sensitivity analyses by removing the studies that violated HWE in the control group, the studies of low quality and the studies that used adult data, indicating that HWE, study quality which included several items such as selection of the study subjects, and age of study participants might be potential sources of heterogeneity. However, because of the limited availability of data from each individual study, the exact source of the heterogeneity could not track, and meta-regression is also not feasible and/or meaningful, again due to the limited number of studies.

For the gene-based and gene-cluster analyses, we adopted the same approach as in our previous work which revealed that, despite weak evidence of individual genetic variants in actotransferrin (*LTF*), multiple genetic variants in *LTF* showed joint contribution to the risk of dental caries^14^. Both of our studies indicated that although the effect of a single genetic variant may be small or insignificant, with proper methods we could still capture the joint contribution of multiple genetic variants. However, our previous study was limited to the exploration of genetic variants within a single gene, whereas in the present study, we examined multiple genetic variants in multiple genes. As a result, we were able to examine a larger number of genetic variants from a larger number of publications, and findings from our gene-based and gene-cluster analysis represented the joint effects from a larger of genetic variants.

Our study has some limitations. Despite efforts in the systematic literature search, the sample sizes for many meta-analyses were limited. As a result, our findings need to be validated by future studies with larger sample sizes. The association of the risk of dental caries with many genetic variants was based on individual studies, which may be subject to biases due to a number of factors such as small sample size and the genetic background of the study participants. The exact relationship of these genetic variants with the risk of dental caries warrant further research. We did not perform analysis by type of dentition because of limited information and limited number of studies. Finally, due to a lack of data for individual subjects, out meta-analyses did not control for important factors that may affect the risk of dental caries. The estimated effect of the reported genetic variants on the risk of dental caries could be greatly confounded by such factors, a limitation of any meta-analysis that uses unadjusted analysis. Future research on the relationship between enamel-formation-related genes and the risk of dental caries should take into account the important confounding factors.

## Conclusions

The present meta-analysis revealed that genetic variant rs17878486 in *AMELX* was associated with dental caries, and multiple genetic variants in enamel-formation-related genes jointly contribute to the risk of dental caries. Future studies with large sample sizes that control for important confounding factors, such as diet, microbial and host characteristics, are needed to validate our findings and to explore additional genetic loci in this gene cluster that might also affect the risk of dental caries.

### Data Availability

No additional data are available.

## Data Availability

No additional data are available.

## Supplementary Materials

Supplementary Table 1-5 provided results for the association of genetic variants in the enamel-formation genes with the risk of dental caries assuming different genetic models.

Supplementary Table 6-8 provided results for sensitivity/subgroup analysis in meta-analyzing four genetic variants in the enamel-formation genes in association with the risk of dental caries.

